# Global surveillance of novel SARS-CoV-2 variants

**DOI:** 10.1101/2023.04.10.23288358

**Authors:** Selina Patel, Fergus Cumming, Carl Mayers, André Charlett, Steven Riley

## Abstract

Earlier global detection of novel SARS-CoV-2 variants gives governments more time to respond. However, few countries can implement timely national surveillance resulting in gaps in monitoring. The UK implemented large-scale community and hospital surveillance, but experience suggests it may be faster to detect new variants through testing UK arrivals for surveillance. We developed simulations of the emergence and importation of novel variants with a range of infection hospitalisation rates (IHR) to the UK. We compared time taken to detect the variant though testing arrivals at UK borders, hospital admissions, and the general community. We found that sampling 10-50% of arrivals at UK borders could confer a speed advantage of 3.5-6 weeks over existing community surveillance, and 1.5–5 weeks (depending on IHR) over hospital testing. We conclude that directing limited global capacity for surveillance to highly connected ports could speed up global detection of novel SARS-CoV-2 variants.

## Introduction

In the current phase of the pandemic, waves of SARS-CoV-2 infection are driven by novel variants and their sub-lineages which continue to cause morbidity and mortality with potential to disrupt society. Government policies to mitigate impacts are more effective if implemented early but have substantial associated costs and therefore should not be implemented unless necessary. Evaluating the threat of an emergent variant to determine a proportionate response requires time to gather evidence. Global surveillance of SARS-CoV-2 and other respiratory pathogen genome sequences aims to contribute to the rapid detection of novel variants so that countries have more time to make policy decisions to respond. However, few countries have capacity and resource to implement timely national surveillance, resulting in gaps in international monitoring.

During the first few years of the pandemic, Hong Kong implemented a strict traveller quarantine protocol.(*1*) Travellers underwent testing for SARS-CoV-2 infection during their quarantine and 10% of detected imported infections were sequenced. Retrospective sequence data from these travellers reflects the global emergence and spread of variants over time. In some cases, traveller-based testing in Hong Kong detected variant circulation in other nations before it had been domestically sequenced and uploaded to the Global Initiative on Sharing Avian Influenza Data (GISAID). The Hong Kong border screening experience suggests that there may be opportunities for traveller-based surveillance to speed up detection of novel variants and compensate for internationally incomplete coverage of domestic genomic surveillance.

To pilot this approach, the US sampled arrival flights from countries with a high travel volume (India, South Africa, Nigeria, Brazil, France, UK, Germany) for voluntary surveillance testing.(*2*) Between November 2021 and January 2022, they achieved a 10% response rate and detected Omicron BA.2 7-days earlier and Omicron BA.3 43-days earlier than anywhere else in the country.

In the UK, although traveller-based surveillance was not implemented when border measures were stepped down in 2022, previous traveller-based testing policies required inbound passengers to undergo testing shortly after arrival.(*3*) The UK also implemented one of the largest community surveys of SARS-CoV-2 surveillance in the world, and all patients experiencing symptomatic respiratory disease in hospital undergo testing for SARS-CoV-2 infection.(*4*) Although reporting times were variable across these testing routes, the first sample from which Omicron was isolated in the England was a mandatory day-2 border test in an inbound traveller on 16^th^ November 2021.(*5*) This was 5-days earlier than the first non-travel associated sample which was obtained on 21^st^ November. Moreover, most of the earliest samples of Delta during the first 2-weeks of detection in the UK were also collected from travellers, despite the availability of universal testing in the community alongside surveillance at that time.(*6*)

To explore the potential utility of border screening for the more rapid detection of variants, we simulate the time to obtaining a sample of an imported novel variant for genomic sequencing through sampling arrivals at ports in England, compared to existing large-scale community surveillance and testing those presenting to hospital. Variants in our scenarios are imported from a country of a similar level of connectedness with England as that currently of China. Over the most recent winter (Dec 2022 – Jan 2023), China experienced a huge increase in transmission of SARS-CoV-2 and resulting deaths following lifting of regulations which were part of previous Zero-covid policy.(7) This risks the emergence of novel variants which could have a significant effect on the epidemiology of COVID-19 elsewhere in the world. We replicate simulations for four scenarios of imported novel variants having an infection hospitalisation rate (IHR) of 1.0%, 1.5%, 2.0% and 2.5%. During the initial spread of Alpha the IHR was estimated between 1.0 and 2.0%, which caused significant impact and resulted in the reintroduction of national lockdown laws to mitigate its spread.(*8,9*)

## Method

A single wave epidemic curve originating in an area with a total population of 60 million was generated. The index case occurred on day 0. A Poisson distribution with a mean of 2 was assumed as the offspring distribution, i.e., each case, on average, transmits an infection to two other people. The distribution of the generation time (the interval between the infection in a primary case and the infection in a secondary case caused by a transmission from the primary case) was assumed to be a Gamma distribution with a shape parameter of 7 and a scale parameter of 1 Thus, the effective reproduction number is 2 and the average doubling time is 7 days. To ensure that the epidemic establishes, the offspring distribution for the first two generations was fixed at exactly two. It was assumed that the epidemic grew unchecked for 16 weeks, after which the mean of the offspring distribution was reduced to represent both control countermeasures and depletion of susceptibles in the population. Between the 17^th^ to the 26^th^ generation, the mean was reduced by 0.1 at each successive generation such that the reproduction number was 1 at the 26^th^ generation. From the 27^th^ generation onwards, the mean of the offspring distribution was reduced at each generation by 0.01786 (1/56).

The incubation period for each generated infection was drawn from the published pooled lognormal distribution in McAloon et al..(*10*) This provides an estimated mean and standard deviation of 1.63 and 0.5 for a Normal distribution of the logged incubation period distribution. Published estimates of the infectious period prior to and post symptom onset are extremely heterogeneous, as described in Byrne et al.(*11*) Thus, the pre-symptomatic infectious period was fixed at 2 days, and the combined pre- and post-symptom infectious period for each generated infection was drawn from a Normal distribution with a mean of 10 days and a standard deviation of 1.33 days. This provides a relatively small probability of being infectious 10 days after symptom onset as found by Singanayagam et al..(*12*) These two periods were rounded to an integer, providing the duration for disease. Daily prevalence is estimated by combining the simulated cases over their duration for all days after the day the index case occurred. In these simulations the period post infectiousness where PCR tests could still detect virus has been ignored. The simulated epidemic curve was truncated at 300 days.

The number of incoming travellers on each day that were incubating or infectious was obtained using a draw from a binomial distribution, with the number of daily travellers assumed to be fixed at 250, and a probability equal to the origin areas prevalence on that day, i.e., assuming that those infected as equally likely to travel as those not infected.

For detection at the border, conditional on the simulations having at least one infected traveller, a representative sample ranging between 10% and 50% of travellers were selected for testing. We further assume that the percentage that are in an infectious state (detectable), the sensitivity of the test, and the percentage of test positives sequenced are 73%, 85% and 50%, respectively. These percentages were used as the probability of draws from independent Bernoulli distributions, with a detection being declared if each of these draws were 1.

Growth in the destination country was assumed to be the same as growth in the origin area as previously described. Incubating or infectious incursions were drawn from a Bernoulli distribution with a probability of 73%. The time remaining in these states was obtained from a uniform distribution, and the mean of the offspring distribution modified to account for this. It was assumed that travellers would spend all of their infectious period in the destination country. Daily incidence and prevalence of cases in the destination country were generated as previously described but with the destination country population being assumed to be 56 million. One thousand destination country epidemics were simulated.

For detection of a simulated case in the hospital setting, IHRs of 1.0% – 2.5% were assumed, and simulated cases were allocated to present at hospital using a draw from a Bernoulli distribution with a probability of 1%. The time to presentation at hospital from infection was assumed to follow a Gamma distribution with a shape parameter of 1.4 and a scale parameter of 4, i.e., giving mean of 5.6 days, but with substantial variation. The percentage of presentations tested was 50%, with the sensitivity of the test, and the percentage of test positives sequenced set as previously stated. Simulations were applied to each of the 1,000 destination country epidemics.

For detection of a simulated case in a community setting, a range of community cohort surveillance sizes between 20,000 (∼0.04% of the population) and 200,000 (∼0.36% of the population) were used. It was assumed each subject in this surveillance was tested every fortnight. Simulations were applied to each of the 1,000 destination country epidemics, with the number detected each day obtained from a draw from a binomial distribution using the number tested each day and the simulated daily prevalence, with the sensitivity of the test, and the percentage of test positives sequenced set as previously.

The time to detecting a case from border, hospital and community testing has been summarised using the empirical 5^th^, 25^th^, 50^th^, 75^th^, and 95^th^ percentile of the simulation sets. For all simulation sets, a unique random number seed was used in a 64-bit Mersenne Twister pseudorandom-number generator. A technical description of the methods is provided in Appendix 1.

## Results

First, we simulated the time to detection of an imported novel variant through different sampling fractions (10%, 20%, 30%, 40%, and 50%) of traveller arrivals in England. We assumed that the prevalence of infection in the passenger population was equal to that of the epidemic curve generated for the country of origin over time (Appendix 1 – Supplementary Figure 4). In our scenarios, there was a non-linear relationship between increasing sampling fraction and decreasing days to detection starting from 131 days to detection through sampling 10% of passenger arrivals (Table 1). The greatest reduction in time to detection was gained between sampling fractions 10 to 20% which led to a median 8-day decrease in time to detection. Thereafter the time gained began to decrease with increasing sampling fraction.

**Table 1.** Simulated time to detect a novel variant since index case through traveller testing for surveillance

| Percentage tested | Empirical centiles of the simulated time to detection distribution (days) |  |  |  |  |
| --- | --- | --- | --- | --- | --- |
|  | 5<sup>th</sup> | 25<sup>th</sup> | **Median** | 75<sup>th</sup> | 95<sup>th</sup> |
| 10 | 104 | 121 | **131** | 140 | 150 |
| 20 | 96 | 114 | **123** | 131 | 141 |
| 30 | 94 | 110 | **119** | 126 | 136 |
| 40 | 89 | 107 | **115** | 123 | 131 |
| 50 | 86 | 105 | **114** | 121 | 130 |

Next, we simulated the time to detection through testing 50% of those presenting to hospital in England. We assumed that growth in incidence in ‘England’ (the destination country) was the same as that in the country of origin. We ran simulations for scenarios where variants had an IHR of 1.0%, 1.5%, 2.0% and 2.5%. Although time to detection in hospitals decreased with increasing IHR, in all four scenarios it took more than 10 days longer to detect a novel variant in hospitals than sampling 10 – 50% of travellers arriving in England (Table 2).

**Table 2.**
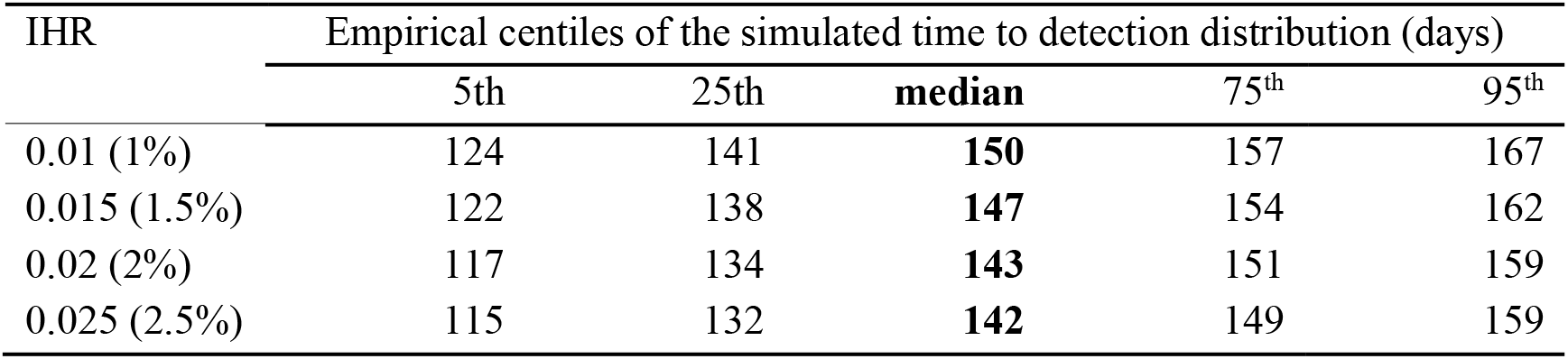
Simulated time to detect a novel variant since index case through hospital testing for surveillance

| IHR | Empirical centiles of the simulated time to detection distribution (days) |  |  |  |  |
| --- | --- | --- | --- | --- | --- |
|  | 5<sup>th</sup> | 25<sup>th</sup> | **median** | 75<sup>th</sup> | 95<sup>th</sup> |
| 0.01 (1%) | 124 | 141 | **150** | 157 | 167 |
| 0.015 (1.5%) | 122 | 138 | **147** | 154 | 162 |
| 0.02 (2%) | 117 | 134 | **143** | 151 | 159 |
| 0.025 (2.5%) | 115 | 132 | **142** | 149 | 159 |

Finally, we simulated the earliest time to obtaining a sample of an imported novel variant through testing a community cohort sampled for surveillance. We ran scenarios implementing a sample size of 0.04 (20,000) – 0.36% (200,000) of the population in England, assuming the same growth in prevalence in the population over time as that assumed for incidence above. Increasing the size of the community cohort from 0.04 to 0.36% of the population, decreased the time to detection by 3 weeks (175 days reduced to 154 days) (Table 3). For the sample size of existing community surveillance in England, which is comprised of around fortnightly 140,000 tests, the simulated earliest time to detection was 157 days.

**Table 3.** Simulated time to detect a novel variant since index case through community testing for surveillance

| Community testing cohort size (% destination country population) | Time to detection (days since emergence of index case) |  |  |  |  |
| --- | --- | --- | --- | --- | --- |
|  | Summaries from 1000 simulations |  |  |  |  |
|  | 5<sup>th</sup> centile | 25<sup>th</sup> centile | **Median** | 75<sup>th</sup> centile | 95<sup>th</sup> centile |
| 20000 (0.04) | 145 | 165 | **175** | 183 | 191 |
| 30000 (0.05) | 144 | 161 | **170** | 178 | 187 |
| 40000 (0.07) | 140 | 158 | **168** | 176 | 185 |
| 50000 (0.09) | 138.5 | 156 | **166** | 175 | 184 |
| 60000 (0.11) | 137 | 155.5 | **165** | 172 | 182 |
| 70000 (0.13) | 137 | 154 | **163** | 171 | 181 |
| 80000 (0.14) | 136 | 153 | **162** | 170 | 179 |
| 90000 (0.16) | 133 | 151 | **161** | 169 | 177 |
| 100000 (0.18) | 133.5 | 150.5 | **160** | 168 | 178 |
| 110000 (0.20) | 130 | 150 | **159** | 167 | 176 |
| 120000 (0.21) | 130 | 148 | **158** | 166.5 | 176 |
| 130000 (0.23) | 130.5 | 149 | **158** | 165 | 174 |
| 140000 (0.25) | 129 | 148 | **157** | 164 | 173 |
| 150000 (0.27) | 129 | 146 | **156** | 163 | 172 |
| 160000 (0.29) | 127.5 | 146 | **155.5** | 163 | 172 |
| 170000 (0.30) | 127 | 146 | **155** | 164 | 173 |
| 180000 (0.32) | 126 | 145 | **154** | 162 | 171 |
| 190000 (0.34) | 128 | 145 | **154** | 162 | 173 |
| 200000 (0.36) | 127 | 144.5 | **154** | 162 | 171 |

We therefore found that, for border testing, the range of the median time to detection from the index case was 131 days (10% tested) to 114 days (50% testing). This compares to 150 days (1% IHR) to 142 days (2.5% IHR) for the median of the earliest time to detection in hospitals, assuming 50% of those presenting are tested. As well as, 175 days (testing a cohort of 0.04% of the population) to 154 days (testing a cohort of 0.36% of the population) for the median of the earliest time to detection through community surveillance.

The complete set of results of this study are provided in Appendix 2.

## Discussion

Our simulations indicate that sampling a relatively small proportion of 10% of inbound travellers for surveillance could reduce the time to detection of the first case of an imported novel variant of SARS-CoV-2 in the England by 26 days, compared to existing community surveillance. Increasing sampling fraction of travellers to 50% could increase this speed advantage to 43 days. Depending on IHR (1.0 – 2.5%), sampling 10% of inbound travellers would also detect a variant between 11 and 19 days faster than testing hospital admissions for surveillance. Whilst sampling 50% of arrivals would lead to detection between 4 and 5 weeks faster than hospital testing.

These simulated results appear concordant with the closest available observed data. In the US, testing 10% of passengers on arrival flights from countries with a high travel volume reported Omicron BA.2 7-days earlier and Omicron BA.3 43-days earlier than anywhere else in the country.(*2*) By comparison in our scenarios, a 10% sampling fraction resulted in detection of a novel variant 1.5-4 weeks sooner than in other settings. Unfortunately, the extent to which further comparisons can be drawn between our results and this experience is limited. The scale of community and healthcare surveillance in the US is significantly smaller than is assumed in our scenarios, and, unlike in our scenarios, US arrivals were required to present a negative test result before departure. Additionally, the time between specimen collection and reporting sequence data can be extremely variable between testing pathways which makes it challenging to observe the speed advantage gained in this example through sampling strategy alone.

Our findings are also broadly in agreement with more distantly related retrospective data from community testing, and policies such as managed quarantine services (MQS) and requirement to test on/shortly after arrival in country. Testing inbound travellers has detected or collected some of the earliest samples of imported novel variants nationally and globally, even during periods when universal testing has been available in the community. In Hong Kong, sequence data were collected for 10% of all infections detected through MQS. Retrospective analysis of these records and external data sources indicate that traveller-based testing was either a good reflection, or an early indicator, of the global emergence and spread of novel variants. For example, Omicron (B1.1.529) was detected in Hong Kong through a sample obtained in a MQS on 13^th^ November 2021(13) which was uploaded to GISAID on 23^rd^ November.(*13*) This triggered UK investigations on 24^th^ November, resulting in government intervention in the UK to delay further introduction and spread.(*14*) The majority of the earliest samples of Omicron subsequently collected in the UK were from people who had been recently exposed to travel.(*5*) It is notable in this example that Omicron samples collected through MQS in Hong Kong were able to be used as prospective evidence for policy decisions because of rapid genomic sequencing of samples and data reporting. In the USA, early samples of Omicron were also collected, frequently from individuals with a history of recent travel exposure, however long lag times from data collection to reporting meant that this was not known until 1^st^ December 2021.(*15*)

We also report that sampling 50% of hospital-presentations for surveillance in our scenarios detects a novel variant with an IHR of 2.5% ∼8 days faster than a variant with an IHR of 1.0%. An increased number of hospital presentations when IHR is greater reduces the speed advantage gained through traveller-based surveillance. However, waves of infection caused by variants with higher IHRs are more likely to be detected earlier in the country of emergence as a result of increasing hospital attendances. This often already offers governments outside of the country of emergence some advanced warning of the impact of a new wave of infection associated with greater morbidity and mortality, despite gaps in global genomic surveillance. The greatest potential impact of early detection through genomic surveillance may therefore be for those variants with an IHR significant enough to cause societal disruption, but low enough that it is slower to identify through hospital admissions.

In order to simulate the time to detection of an imported novel variant in England in each of our scenarios, we have made some simplifying assumptions. We have assumed that the prevalence of infection in air passengers is the same as that in the country of origin at the time of the departure of their flight, specimens are collected from a random sample of passengers, and the variant doubling time in the destination country is the same as that of in country of origin once seeded. We have also considered only direct incursions from the country of emergence of a novel variant to the destination country. We have not considered the effect of indirect incursions linked to infected travellers arriving from other countries where transmission may also be occurring. This is a simplification of observed human behaviour, population immunity profiles and transmission dynamics. However, we do not expect that a model comprised of more complex representations of these processes would result in significantly different overall conclusions.

In this report we have focused the results and discussion on simulated scenarios which compare border surveillance with existing surveillance in hospitals and the community in England and the UK. However, this surveillance in England achieved greater coverage than in most countries in the world today. Therefore, as routine testing and surveillance for SARS-CoV-2 is globally wound down, this work likely provides conservative estimates of the potential speed advantage which could be gained through traveller-based surveillance approaches. Also, if there were concerns about a specific country at any point in time, temporary programmes would be able to achieve high sample proportions at the border with only limited numbers of samples compared to other ongoing or potential global programmes.

It is also important to recognise that the collection of a sample of a novel variant for detection is the first step to evaluate the threat of a novel variant. In our scenarios, we do not consider the time it takes to sequence and report data obtained from a sample. Sequencing and reporting times are extremely variable across countries which can significantly reduce the time gained through effective sampling approaches.(*16*) Additionally, a full threat assessment relies on additional data, such as case and hospitalisation patterns and genomic data from community and healthcare surveillance, to gather evidence to inform policy.

Global surveillance of SARS-CoV-2 genome sequences contributes to rapid detection of novel variants to give governments more time to respond. However, few countries have capacity to implement national surveillance with timely sequencing and reporting, resulting in significant gaps in global coverage of surveillance. In our scenarios, directing limited global capacity for surveillance to the most highly connected ports could provide governments with significantly more time to respond to future novel variants of SARS-CoV-2 and their sub-lineages. Beyond informing national approaches to surveillance, this also underscores the potential usefulness of international collaboration to achieve high global coverage of surveillance and provide governments with more time to make policy decisions to respond to novel variants of SARS-CoV-2.

## Supporting information

Appendix_1_technical_method

Appendix_2_results_of_simulations

## Data Availability

All data produced in the present study are provided in Appendix 2.

## Acknowledgements

No additional funding was provided for this work.

## Biographical sketch

Selina Patel is a PhD researcher at University College London and an honorary public health analyst at the UK Health Security Agency. Selina’s research interests span public health related to infectious disease, with a particular focus on surveillance approaches.

## Supplemental Materials

Appendix 1: .docx containing technical description of the method

Appendix 2: .docx containing complete results

