## Appendix_1_technical_method for "Global surveillance of novel SARS-CoV-2 variants"

### Appendix 1: Methodology for earliest detection a novel variant via border testing, hospital, and community surveillance.

Parameters

| Parameter | | Estimates/distributions used |
|---|---|---|
| Population of origin | $N_O$ | 60,000,000 |
| Population of destination | $N_D$ | 56,000,000 |
| Offspring | $c_k$ | $Poisson(2)$ |
| Generation time | $g_k$ | $round(\Gamma(7,1), 1)$ |
| Incubation period | $l_k$ | $round\left(e^{N(1.63,0.5)}, 1\right)$ |
| Infectiousness period | $i_k$ | $round(N(10,1.33), 1)$ |
| Day of infection for the $k^{th}$ simulated infection | $d_k$ | |
| Day of symptom onset for the $k^{th}$ simulated infection | $d_k^s$ | |
| Infection incidence in origin on day $d$ | $IO_d$ | |
| Disease incidence in destination on day $d$ | $ID_d$ | |
| Infection prevalence in origin on day $d$ | $PO_d$ | |
| Disease prevalence in destination on day $d$ | $PD_d$ | |
| Direct travellers per day | $n_t$ | 100, 250, 500 |
| Proportion of travellers tested | $\pi_a$ | 0.01, 0.02, 0.05.0.10, 0.20 |
| Probability infectious $\overline{\iota_k/(l_k + \iota_k - 2)}$ | $\pi_{inf}$ | 0.73 |
| PCR Test sensitivity | $\pi_{sens}$ | 0.85 |
| Positive successfully sequenced | $\pi_{seq}$ | 0.5 |
| IHR | $\pi_{IHR}$ | 0.005,0.01,0.015,0.02,0.025 |
| Proportion of hospital presentation testing | $\pi_h$ | 0.1, 0.2, 0.3, 0.4, 0.5 |
| Time to hospital presentation | $t_h$ | $round((\Gamma(5,2), 1)$ |
| Size of community surveillance | $n_c$ | 20,000 – 200,000 (steps of 10,000) |

<u>Simulated epidemic at origin</u>

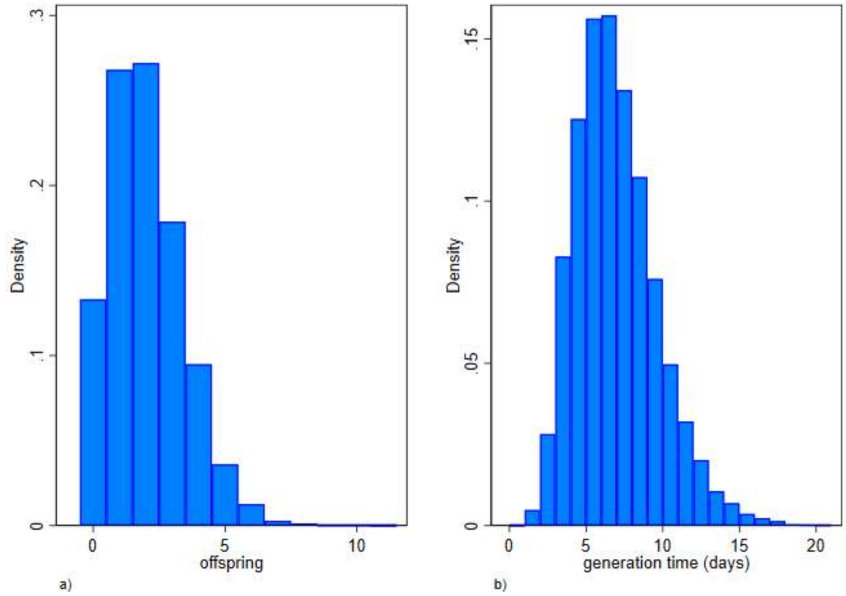


*Supplementary Figure 1: offspring (panel a) and generation time (panel b) distributions*

A single occurrence of a novel variant occurs on day zero $(d_0)$. To obtain the resultant epidemic curve, offspring, and generation time, distributions of $Poisson(2)$ and $\Gamma(7,1)$ were used respectively with the offspring fixed a 2 for the first two generations to ensure the epidemic establishes. Random draws of generation times were rounded to the nearest integer.

This choice of offspring and generation time provides an epidemic curve where $R_0$ is 2 with a doubling time of 7 days. The variant was assumed to grow unchecked for 16 generations, after which the mean of the offspring distribution was reduced by 0.1 of each successive generation to obtain an offspring distribution $Poisson(1)$ at the 26$^{th}$ generation, i.e., $R_t$ of 1 after around 6 months. From the 27$^{th}$ generation onwards, the mean of the offspring distribution was reduced at each generation by 0.01786 (1/56).

Fifty-two generations were simulated, but a 300-day cut off used to ensure that chains with shorter than average generation times did not impact completeness of simulated infections, there being a probability of around 0.0002 of obtaining a sum of 52 draws from $round(\Gamma(7,1), 1)$ being less than 300.

For each simulated infection $k$, the day of infection $d_k$ was obtained by summation of the generation times for their predecessors. The incidence on day $d$ is obtained via a summation over all $n$ infections, $IO_d = \sum_{k=1}^{n}(d_k == d)$, where $(d_k == d)$ is 1 if $d_k$ equals $d$, 0 otherwise. The simulated epidemic curve is shown in Supplementary Figure 2, for which the cumulative incidence up to day 300 since $d_0$ is around 23,000,000.

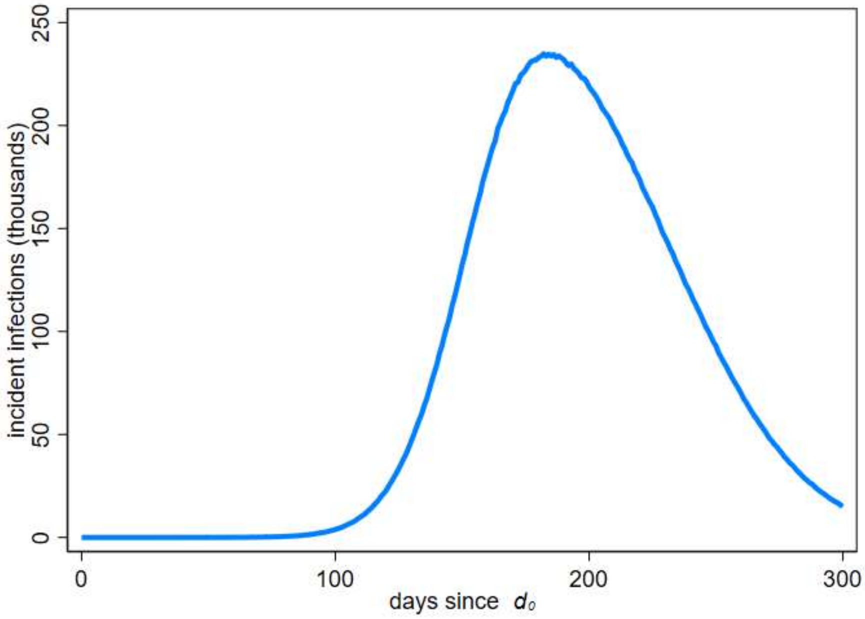


*Supplementary Figure 2: Simulated epidemic curve at the origin.*

For each simulated infection, a log incubation period was obtained from a random draw from $N(1.63,0.5)$, with time of symptom onset occurring at $d_k^s = d_k + l_k$. The 10$^{th}$, 50$^{th}$, and 90$^{th}$ centiles for this incubation period distribution are approximately 2.7, 5.1, and 9.7 days, respectively.

The infectious period distribution was assumed to be $N(10,1.33)$, with the period of infectiousness beginning two days prior to symptom onset. Thus, the first day of being infectious for the $k^{th}$ infection occurs on day $d_k^s - 2$. If $d_k^s - 2 < d_k$ then $l_k$ is set to 0, and if $d_k^s - 2 > 19$ then $l_k$ is set to 19. Supplementary Figure 3 provides a visualisation of the assumed incubation period distribution and the temporal probability of infectiousness.

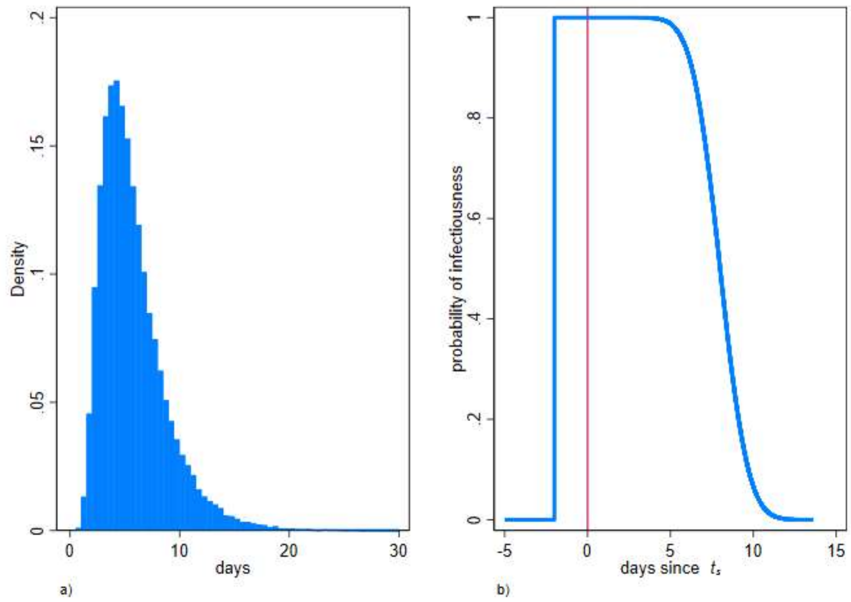


*Supplementary Figure 3: incubation period distribution (panel a) and the probability of infectiousness (panel b)*

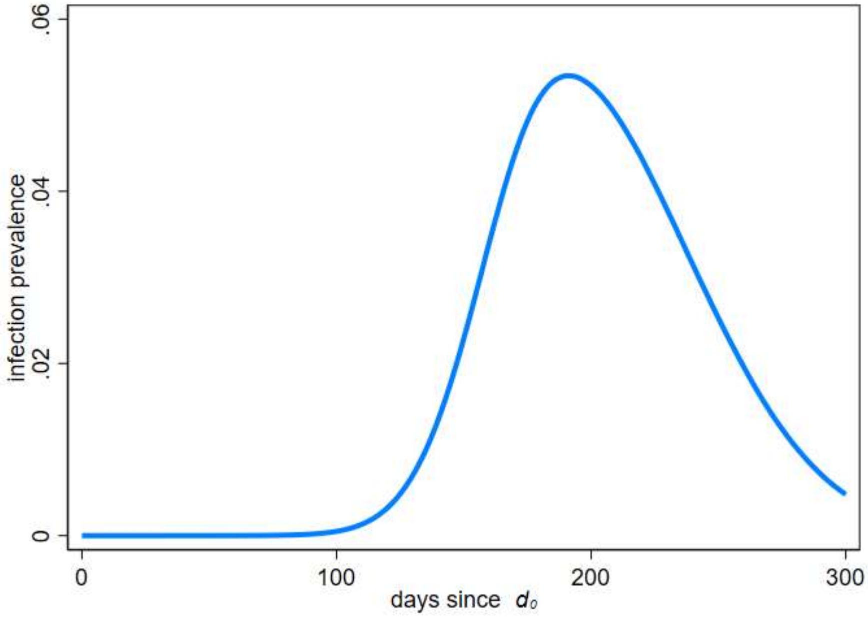


*Supplementary Figure 4: Simulated infection prevalence at the origin*

The pre-infectious period ($d_k$ to $d_k^s - 3$) and infectious period ($d_k^s - 2$ to $d_k^s - 2 + i_k$) are combined and the prevalence of being in either state on day $d$ is obtained from $PO_d = \frac{1}{N_o}\sum_{k=1}^{n}\left(d_k \leq d \leq (d_k + l_k + i_k - 2)\right)$, where $d_k \leq d \leq (d_k + l_k + i_k - 2)$ is 1 if $d$ is greater or equal to $d_k$and less than or equal to $d_k + l_k + i_k - 2$, 0 otherwise. The infection prevalence is shown in Supplementary Figure 4.

Simulated epidemics at destination

For each day from $d_0$ it is assumed for simplicity, that there are a fixed number of direct air travellers $n_t$ departing the origin for the destination. These travellers are assumed to have the same infection prevalence as the origin on the day of departure and all flights depart and arrive on the same day. On day $d$ the number of infected travellers is obtained from a random draw from $binomial(n_t, PO_d)$. A simulated infected traveller is in the infectious state if a random draw from $binomial(1, \pi_{inf})$ is equal to 1, otherwise they are in a pre-infectious state. For the simulated incursions in both the pre-infectious and infectious state, the time in their state has been reduced by multiplication with a random draw from a uniform distribution $Uniform([0,1])$, and for those in the infectious state, their simulated number of offspring has similarly been reduced.

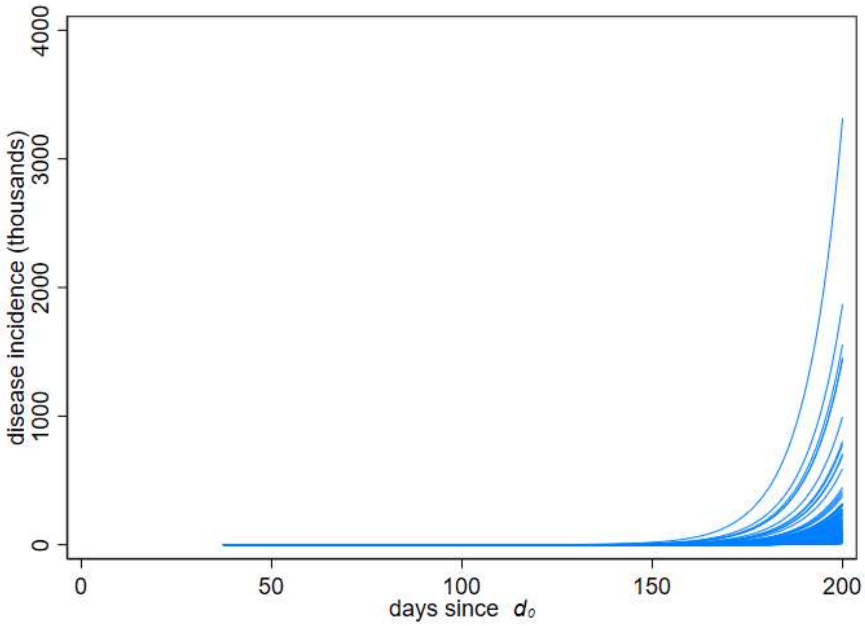


*Supplementary Figure 5: Simulated disease incidence growth curves within the destination country for 500 arrivals per day using daily prevalence from Supplementary Figure 4*

One thousand simulated incidence growth curves have been generated from the simulated incursions, for a total of 30 generations. For simplicity, it is assumed that the offspring distribution and generation time distribution are the same as at the origin. While this is likely to be true for the latter, for the former this would implicitly assume that population mixing in the origin and destination are similar. It has also been assumed that detection is only possible during the infectious state, and the post infectiousness period where PCR tests could still detect virus has been ignored. Thus, $ID_d = \sum_{k=1}^{n}(d_k^s == d)$, where $(d_k^s == d)$ is 1 if $d_k^s$ equals $d$, 0 otherwise.

The incidence growth curves have been converted to disease prevalence growth curves using the same methodology previously described but ignoring the days in the pre-infectious state. These are shown in Figures 5 and 6

The disease prevalence on day $d$ is obtained from $PD_d = \frac{1}{N_D}\sum_{k=1}^{n}\left(d_k^s - 2 \leq d \ \leq \ (d_k^s - 2 + i_k)\right)$, where $d_k^s - 2 \leq d \ \leq \ (d_k^s - 2 + i_k)$ is 1 if $d$ is greater or equal to $d_k^s - 2$ and less than or equal to $d_k^s - 2 + i_k$, 0 otherwise.

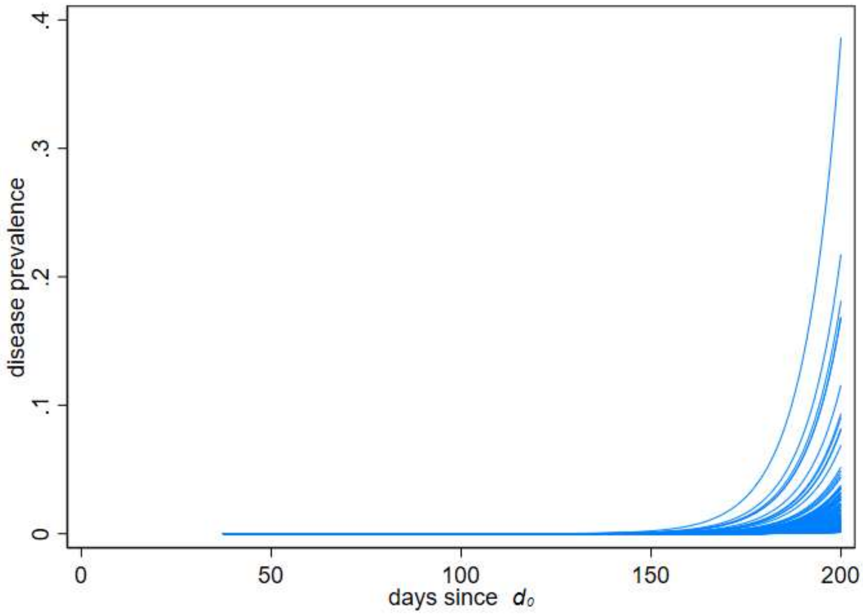


*Supplementary Figure 6: Simulated disease prevalence growth curves within the destination country for 500 arrivals per day*

While for most simulations the growth of incident cases is modest, there are a small proportion where exceptional growth is observed. As the objective was to gain an understanding of the earliest time to detection, no account has been taken of either depletion of those susceptible in the population or effective control measures both of which would cause the simulations with exceptional growth to turn over and decline. It has been assumed that such behaviour would only occur post the earliest detections so are not an important consideration.

<u>Time to earliest detection</u>

For each of testing arrivals at the border, testing those sufficiently ill to present at hospital, and testing those enrolled in community surveillance, simulations were implemented as outlined in future sections with one thousand such simulations performed for each. For all simulation sets, a unique random-number seed was used in a 64-bit Mersenne Twister pseudo-random-number generator.

The simulated earliest time of detection, i.e., earliest specimen date, ignoring any sample processing and reporting delays is obtained for each simulation with selected centiles of the simulated distributions of earliest detection times presented. For results in the paper, the median time to earliest detection is presented, with time in relation to the occurrence of the very first case $d_0$ being used throughout.

Time to earliest detection for testing at the border $(T_b)$

The number of incoming travellers $n_d^I$ on each day that are either incubating or infectious is obtained using a draw from $binomial(n_t, PO_d)$. For simplicity, an assumption that those travelling are independent of infection status has been made. Whilst this assumption will influence the time to earliest detection, which would occur later if prevalence in travellers is lower, it is unlikely to impact greatly any relative difference in the times to earliest detection at the border, or in either hospital or community surveillance.

The number of infected travellers being tested on day $d$, $n_d^{tested}$is obtained using a draw from $binomial\left(n_d^I, \pi_t\right)$, provided $n_d^I$>0. The test on the $k^{th}$ simulated infection is considered positive $(b_k^+)$ if it is an infectious state, not having a false negative test, and being successfully sequenced. This was obtained by multiplying the random draws from three Bernoulli distributions, $binomial\left(1, \pi_{inf}\right)$, $binomial(1, \pi_{sens})$, and $binomial\left(1, \pi_{seq}\right)$, with a detection being declared if each of these draws are 1, otherwise considered a failure to detect. It has been assumed that the microbiological test has a sensitivity of 85%, and specificity of 100%, and that for technical reasons only 50% of positives isolates will lead to a sequence being successfully obtained.

For each detection at the border $b_k^+$, $d_k^{b^+}$is the day on which the positive specimen is taken. The time to earliest detection is $T_b = \min_{k \in \{b_k^+\}} d_k^{b^+}$.

Time to earliest detection for testing at hospitals $(T_h)$

From each of the 1000 simulated disease incidence growth curves in the destination country, the number of infections each day that would result in a hospital admission $n_d^h$ was obtained from a random draw from $binomial(ID_d, \pi_{IHR})$, provided $ID_d$>0. The number of those hospitalised that will get tested, $n_d^{h^t}$ was obtained from a random draw from $binomial\left(n_d^h, \pi_h\right)$, provided $n_d^h$>0.

The $k^{th}$ infected simulation is considered positive $h_k^+$ if it is not a false negative test and is successfully sequenced. This was obtained by multiplying random draws from two Bernoulli distributions, $binomial(1, \pi_{sens})$, and $binomial\left(1, \pi_{seq}\right)$, with a detection being declared if each of both these draws are 1, otherwise considered a failure to detect.

For each hospital detection $h_k^+$, the day on which the positive specimen is taken $d_k^{h^+}$ was obtained from $d_k^s + t_h$, where $d_k^s$ is the day of symptom onset of the $k^{th}$ infection, and $t_h$ is the time from symptom onset to hospitalisation, taken as a random draw from a $\Gamma(5,2)$ and rounded to the nearest integer as shown in Supplementary Figure 7. The time to earliest detection is obtained using $T_h = \min_{k \in \{h_k^+\}} d_k^{h^+}$.

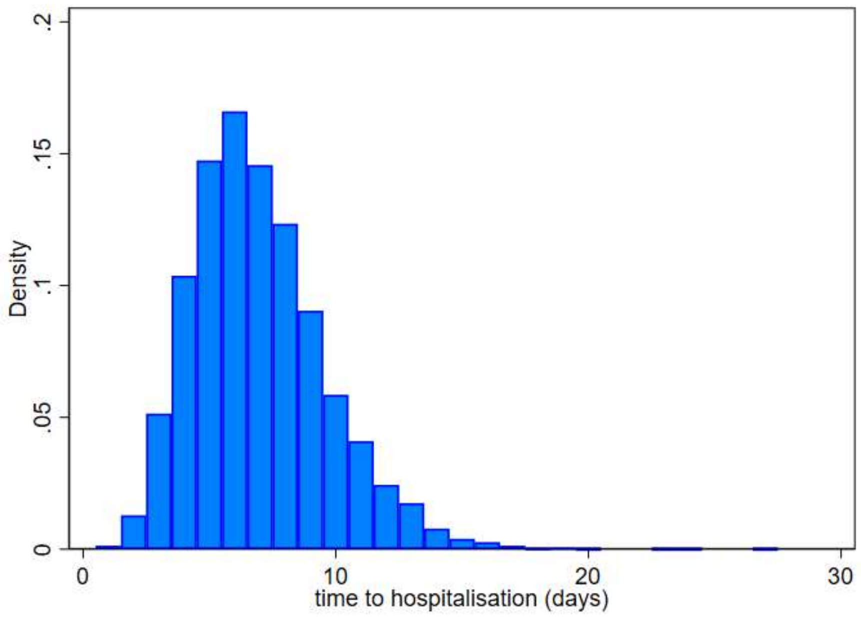


*Supplementary Figure 7: Time from symptom onset to hospitalisation distribution*

Time to earliest detection for testing in the community ($T_c$)

It has been assumed that individuals in community surveillance test every fortnight, and for simplicity the number of daily tests is $\frac{n_c}{14}$. From each of the 1000 simulated disease prevalence growth curves in the destination country, the number of simulated prevalent infections that would be tested on day $d$ in community surveillance $n_d^{c^t}$ was obtained from a random draw from $binomial\left(round\left(\frac{n_c}{14},1\right),PD_d\right)$, provided $PD_d$>0.

The $k^{th}$ infected simulation is considered positive $c_k^+$ if it is not a false negative test and is successfully sequenced. As before this was obtained by multiplying random draws from two Bernoulli distributions, $binomial(1,\pi_{sens})$, and $binomial(1,\pi_{seq})$, with a detection being declared if each of these draws are 1, otherwise considered a failure to detect. For each detection in the community $c_k^+$, $d_k^{c^+}$ is the day on which the positive specimen is taken. The time to earliest detection is obtained using $T_c = \min_{k\in\{c_k^+\}} d_k^{c^+}$.
