## Appendix_2_results_of_simulations for "Global surveillance of novel SARS-CoV-2 variants"

The tables below provide the results of the simulations described in the main paper and Appendix 1. The nature of simulations means that each time they are run, there will be slight variation in the results produced (not significant enough to change the trends or overall conclusions reported).

Table B1: Summary statistics for the simulated earliest time to detection distribution for testing at the border with 250 daily passengers, all times refer to days since the index case

| Proportion tested | centiles | | | | | | | | |
|---|---|---|---|---|---|---|---|---|---|
| | 1st | 5th | 10th | 25th | 50th | 75th | 90th | 95th | 99th |
| 0.010 | 115 | 127 | 136 | 150 | 166 | 184 | 206 | 221 | 253 |
| 0.020 | 106 | 120 | 127 | 140 | 152 | 168 | 181 | 189 | 206 |
| 0.030 | 98 | 112 | 121 | 133 | 146 | 158 | 169 | 174 | 189 |
| 0.040 | 95 | 108 | 118 | 131 | 143 | 154 | 162 | 167 | 178 |
| 0.050 | 91 | 111 | 118 | 128 | 139 | 149 | 157 | 162 | 175 |
| 0.060 | 95 | 107 | 116 | 125 | 137 | 146 | 155 | 161 | 171 |
| 0.070 | 90 | 109 | 116 | 125 | 135 | 144 | 152 | 156 | 164 |
| 0.080 | 94 | 106 | 114 | 124 | 135 | 143 | 151 | 155 | 162 |
| 0.090 | 92 | 106 | 112 | 121 | 131 | 140 | 149 | 153 | 160 |
| 0.100 | 90 | 104 | 109 | 121 | 131 | 140 | 147 | 150 | 157 |
| 0.110 | 89 | 100 | 109 | 120 | 130 | 139 | 146 | 150 | 158 |
| 0.120 | 88 | 101 | 109 | 119 | 128 | 137 | 143 | 147 | 156 |
| 0.130 | 88 | 102 | 108 | 119 | 128 | 136 | 143 | 148 | 153 |
| 0.140 | 86 | 96 | 105 | 116 | 127 | 135 | 141 | 144 | 151 |
| 0.150 | 80 | 99 | 106 | 116 | 126 | 135 | 141 | 145 | 151 |
| 0.160 | 81 | 99 | 105 | 116 | 126 | 134 | 141 | 144 | 151 |
| 0.170 | 84 | 98 | 106 | 115 | 123 | 132 | 140 | 144 | 148 |
| 0.180 | 80 | 97 | 105 | 115 | 124 | 132 | 138 | 142 | 149 |
| 0.190 | 79 | 98 | 106 | 115 | 123 | 132 | 138 | 141 | 147 |
| 0.200 | 85 | 96 | 104 | 114 | 123 | 131 | 137 | 141 | 147 |
| 0.210 | 80 | 97 | 104 | 113 | 123 | 130 | 137 | 140 | 146 |
| 0.220 | 81 | 96 | 102 | 112 | 122 | 129 | 135 | 139 | 145 |
| 0.230 | 78 | 96 | 103 | 111 | 121 | 129 | 135 | 138 | 144 |
| 0.240 | 76 | 92 | 100 | 110 | 120 | 128 | 135 | 138 | 145 |
| 0.250 | 80 | 94 | 100 | 110 | 120 | 128 | 134 | 137 | 143 |
| 0.260 | 82 | 97 | 101 | 110 | 120 | 126 | 133 | 136 | 142 |
| 0.270 | 76 | 95 | 102 | 110 | 119 | 127 | 133 | 137 | 144 |
| 0.280 | 78 | 95 | 101 | 111 | 119 | 126 | 132 | 136 | 142 |
| 0.290 | 73 | 92 | 101 | 109 | 118 | 126 | 132 | 136 | 142 |
| 0.300 | 83 | 94 | 100 | 110 | 119 | 126 | 132 | 136 | 142 |
| 0.310 | 76 | 94 | 100 | 109 | 118 | 126 | 131 | 135 | 141 |
| 0.320 | 70 | 90 | 98 | 109 | 117 | 125 | 131 | 134 | 140 |
| 0.330 | 72 | 89 | 97 | 108 | 117 | 125 | 131 | 135 | 140 |
| 0.340 | 77 | 92 | 98 | 106 | 117 | 124 | 130 | 133 | 138 |
| 0.350 | 75 | 89 | 97 | 108 | 116 | 124 | 130 | 134 | 139 |
| 0.360 | 72 | 91 | 98 | 107 | 116 | 124 | 129 | 133 | 140 |
| 0.370 | 65 | 90 | 97 | 106 | 115 | 123 | 130 | 133 | 137 |
| 0.380 | 78 | 91 | 97 | 107 | 116 | 124 | 129 | 133 | 138 |
| 0.390 | 66 | 91 | 97 | 106 | 115 | 122 | 129 | 132 | 138 |

| | | | | | | | | | |
|---|---|---|---|---|---|---|---|---|---|
| 0.400 | 78 | 89 | 96 | 107 | 115 | 123 | 128 | 131 | 138 |
| 0.410 | 78 | 90 | 97 | 105 | 115 | 123 | 128 | 131 | 138 |
| 0.420 | 76 | 90 | 97 | 106 | 115 | 122 | 128 | 131 | 136 |
| 0.430 | 74 | 86 | 96 | 107 | 115 | 122 | 128 | 131 | 137 |
| 0.440 | 77 | 90 | 98 | 107 | 115 | 122 | 128 | 130 | 135 |
| 0.450 | 65 | 87 | 95 | 106 | 115 | 122 | 127 | 132 | 138 |
| 0.460 | 68 | 88 | 94 | 105 | 114 | 121 | 126 | 129 | 136 |
| 0.470 | 75 | 90 | 96 | 105 | 113 | 120 | 127 | 129 | 135 |
| 0.480 | 78 | 90 | 96 | 104 | 113 | 120 | 126 | 130 | 135 |
| 0.490 | 73 | 88 | 95 | 105 | 114 | 121 | 126 | 129 | 134 |
| 0.500 | 72 | 86 | 94 | 105 | 114 | 121 | 126 | 130 | 134 |

Table H1: Summary statistics for the simulated earliest time to detection distribution for testing at hospitals with seedings from 250 daily passengers

| IHR | Proportion tested | centiles | | | | | | | | |
|---|---|---|---|---|---|---|---|---|---|---|
| | | 1st | 5th | 10th | 25th | 50th | 75th | 90th | 95th | 99th |
| 0.005 | 0.10 | 128 | 143 | 150 | 161 | 171 | 178 | 185 | 188 | 195 |
| 0.005 | 0.20 | 119 | 139 | 145 | 155 | 164 | 173 | 179 | 181 | 187 |
| 0.005 | 0.30 | 114 | 134 | 140 | 151 | 161 | 168 | 175 | 178 | 183 |
| 0.005 | 0.40 | 116 | 132 | 140 | 150 | 159 | 166 | 172 | 175 | 181 |
| 0.005 | 0.50 | 117 | 128 | 137 | 146 | 156 | 164 | 169 | 173 | 179 |
| 0.010 | 0.10 | 123 | 137 | 144 | 155 | 165 | 172 | 178 | 182 | 187 |
| 0.010 | 0.20 | 115 | 131 | 137 | 147 | 157 | 165 | 172 | 175 | 181 |
| 0.010 | 0.30 | 115 | 129 | 136 | 145 | 154 | 162 | 169 | 172 | 178 |
| 0.010 | 0.40 | 108 | 124 | 131 | 141 | 151 | 159 | 165 | 168 | 173 |
| 0.010 | 0.50 | 107 | 124 | 132 | 141 | 150 | 157 | 164 | 167 | 170 |
| 0.015 | 0.10 | 112 | 133 | 139 | 151 | 161 | 168 | 174 | 178 | 183 |
| 0.015 | 0.20 | 108 | 125 | 133 | 144 | 155 | 162 | 169 | 172 | 177 |
| 0.015 | 0.30 | 101 | 122 | 129 | 142 | 152 | 159 | 165 | 168 | 174 |
| 0.015 | 0.40 | 104 | 120 | 128 | 139 | 147 | 156 | 162 | 165 | 169 |
| 0.015 | 0.50 | 108 | 122 | 128 | 138 | 147 | 154 | 159 | 162 | 167 |
| 0.020 | 0.10 | 115 | 132 | 138 | 149 | 158 | 166 | 171 | 174 | 181 |
| 0.020 | 0.20 | 108 | 123 | 131 | 142 | 152 | 160 | 166 | 168 | 174 |
| 0.020 | 0.30 | 110 | 123 | 128 | 138 | 148 | 156 | 161 | 165 | 172 |
| 0.020 | 0.40 | 102 | 119 | 126 | 137 | 146 | 154 | 160 | 163 | 166 |
| 0.020 | 0.50 | 103 | 117 | 124 | 134 | 143 | 151 | 156 | 159 | 164 |
| 0.025 | 0.10 | 111 | 128 | 135 | 147 | 156 | 163 | 169 | 173 | 177 |
| 0.025 | 0.20 | 108 | 122 | 129 | 140 | 149 | 157 | 162 | 166 | 172 |
| 0.025 | 0.30 | 104 | 118 | 127 | 138 | 147 | 154 | 159 | 162 | 168 |
| 0.025 | 0.40 | 104 | 119 | 125 | 135 | 143 | 151 | 157 | 159 | 164 |
| 0.025 | 0.50 | 104 | 115 | 122 | 132 | 142 | 149 | 155 | 159 | 163 |

Table B2: Summary statistics for the simulated earliest time to detection distribution for testing at the border with 500 daily passengers, all times refer to days since the index case

| Proportion tested | centiles | | | | | | | | |
|---|---|---|---|---|---|---|---|---|---|
| | 1st | 5th | 10th | 25th | 50th | 75th | 90th | 95th | 99th |
| 0.010 | 108 | 124 | 130 | 142 | 154 | 167 | 179 | 186 | 207 |
| 0.020 | 103 | 114 | 122 | 131 | 143 | 153 | 163 | 170 | 181 |
| 0.030 | 94 | 109 | 116 | 127 | 138 | 147 | 156 | 160 | 172 |
| 0.040 | 93 | 106 | 112 | 122 | 135 | 144 | 152 | 156 | 163 |
| 0.050 | 89 | 103 | 111 | 122 | 131 | 140 | 147 | 151 | 161 |
| 0.060 | 87 | 102 | 109 | 118 | 129 | 136 | 143 | 146 | 155 |
| 0.070 | 88 | 100 | 108 | 118 | 127 | 136 | 142 | 146 | 154 |
| 0.080 | 79 | 98 | 106 | 116 | 125 | 133 | 140 | 143 | 149 |
| 0.090 | 84 | 97 | 104 | 114 | 124 | 132 | 138 | 142 | 147 |
| 0.100 | 82 | 96 | 103 | 114 | 124 | 132 | 137 | 140 | 147 |
| 0.110 | 81 | 95 | 103 | 112 | 122 | 130 | 136 | 140 | 146 |
| 0.120 | 79 | 94 | 103 | 112 | 121 | 129 | 136 | 138 | 143 |
| 0.130 | 75 | 93 | 102 | 111 | 120 | 128 | 133 | 137 | 143 |
| 0.140 | 77 | 93 | 100 | 110 | 119 | 127 | 133 | 137 | 140 |
| 0.150 | 79 | 93 | 99 | 109 | 118 | 126 | 132 | 135 | 140 |
| 0.160 | 75 | 92 | 98 | 108 | 118 | 125 | 132 | 136 | 140 |
| 0.170 | 79 | 93 | 99 | 109 | 118 | 125 | 131 | 134 | 140 |
| 0.180 | 73 | 89 | 97 | 108 | 116 | 124 | 130 | 133 | 138 |
| 0.190 | 73 | 88 | 97 | 107 | 115 | 123 | 129 | 133 | 138 |
| 0.200 | 76 | 91 | 97 | 105 | 114 | 122 | 127 | 131 | 137 |
| 0.210 | 79 | 90 | 97 | 106 | 115 | 123 | 129 | 131 | 137 |
| 0.220 | 74 | 92 | 97 | 106 | 114 | 121 | 127 | 130 | 137 |
| 0.230 | 76 | 89 | 96 | 106 | 114 | 121 | 127 | 130 | 136 |
| 0.240 | 72 | 87 | 93 | 104 | 113 | 121 | 126 | 130 | 134 |
| 0.250 | 73 | 88 | 97 | 105 | 113 | 120 | 126 | 129 | 135 |
| 0.260 | 68 | 89 | 95 | 105 | 113 | 120 | 126 | 129 | 133 |
| 0.270 | 71 | 87 | 93 | 103 | 112 | 119 | 125 | 127 | 132 |
| 0.280 | 70 | 87 | 95 | 104 | 112 | 119 | 124 | 128 | 134 |
| 0.290 | 64 | 85 | 92 | 101 | 111 | 119 | 125 | 128 | 133 |
| 0.300 | 65 | 83 | 91 | 102 | 111 | 118 | 125 | 127 | 132 |
| 0.310 | 70 | 85 | 92 | 103 | 111 | 118 | 123 | 126 | 131 |
| 0.320 | 70 | 86 | 92 | 102 | 111 | 117 | 123 | 125 | 130 |
| 0.330 | 68 | 86 | 92 | 101 | 110 | 117 | 124 | 127 | 131 |
| 0.340 | 71 | 84 | 90 | 101 | 110 | 117 | 123 | 126 | 131 |
| 0.350 | 71 | 87 | 93 | 101 | 111 | 117 | 123 | 125 | 131 |
| 0.360 | 69 | 86 | 92 | 100 | 109 | 116 | 121 | 125 | 130 |
| 0.370 | 68 | 84 | 91 | 101 | 109 | 116 | 122 | 125 | 129 |
| 0.380 | 68 | 84 | 92 | 100 | 109 | 116 | 121 | 124 | 127 |
| 0.390 | 68 | 84 | 89 | 101 | 109 | 116 | 120 | 123 | 128 |
| 0.400 | 71 | 84 | 90 | 100 | 109 | 116 | 121 | 123 | 128 |
| 0.410 | 65 | 84 | 90 | 100 | 108 | 115 | 120 | 123 | 127 |
| 0.420 | 71 | 86 | 91 | 100 | 107 | 114 | 120 | 122 | 127 |
| 0.430 | 72 | 84 | 90 | 99 | 108 | 115 | 121 | 123 | 128 |
| 0.440 | 69 | 82 | 88 | 98 | 108 | 115 | 120 | 123 | 129 |
| 0.450 | 68 | 82 | 88 | 99 | 107 | 114 | 119 | 122 | 127 |
| 0.460 | 65 | 81 | 88 | 98 | 107 | 115 | 120 | 123 | 128 |

| | | | | | | | | | |
|---|---|---|---|---|---|---|---|---|---|
| 0.470 | 56 | 82 | 87 | 98 | 106 | 114 | 119 | 122 | 127 |
| 0.480 | 61 | 83 | 89 | 98 | 106 | 113 | 118 | 121 | 127 |
| 0.490 | 63 | 81 | 88 | 98 | 106 | 113 | 119 | 121 | 125 |
| 0.500 | 63 | 79 | 87 | 97 | 106 | 113 | 118 | 121 | 127 |

Table H2: Summary statistics for the simulated earliest time to detection distribution for testing at hospitals with seedings from 500 daily passengers

| IHR | Proportion tested | centiles | | | | | | | | |
|---|---|---|---|---|---|---|---|---|---|---|
| | | 1st | 5th | 10th | 25th | 50th | 75th | 90th | 95th | 99th |
| 0.005 | 0.10 | 115 | 135 | 143 | 154 | 163 | 171 | 177 | 179 | 184 |
| 0.005 | 0.20 | 114 | 128 | 135 | 147 | 156 | 164 | 169 | 172 | 178 |
| 0.005 | 0.30 | 117 | 127 | 133 | 144 | 153 | 161 | 167 | 169 | 173 |
| 0.005 | 0.40 | 108 | 122 | 131 | 141 | 149 | 157 | 163 | 166 | 172 |
| 0.005 | 0.50 | 109 | 121 | 130 | 140 | 148 | 156 | 161 | 164 | 169 |
| 0.010 | 0.10 | 121 | 132 | 138 | 147 | 156 | 164 | 170 | 174 | 177 |
| 0.010 | 0.20 | 109 | 123 | 128 | 140 | 149 | 157 | 163 | 167 | 172 |
| 0.010 | 0.30 | 104 | 121 | 128 | 137 | 147 | 154 | 160 | 163 | 167 |
| 0.010 | 0.40 | 105 | 119 | 125 | 135 | 144 | 151 | 157 | 160 | 167 |
| 0.010 | 0.50 | 98 | 116 | 123 | 134 | 142 | 149 | 155 | 158 | 163 |
| 0.015 | 0.10 | 110 | 128 | 133 | 144 | 153 | 160 | 165 | 168 | 172 |
| 0.015 | 0.20 | 104 | 120 | 126 | 137 | 147 | 154 | 160 | 162 | 169 |
| 0.015 | 0.30 | 103 | 118 | 125 | 134 | 143 | 151 | 156 | 159 | 164 |
| 0.015 | 0.40 | 100 | 112 | 121 | 132 | 141 | 148 | 154 | 157 | 162 |
| 0.015 | 0.50 | 95 | 112 | 119 | 130 | 139 | 145 | 151 | 154 | 158 |
| 0.020 | 0.10 | 107 | 122 | 130 | 140 | 150 | 158 | 163 | 166 | 172 |
| 0.020 | 0.20 | 106 | 118 | 125 | 135 | 144 | 152 | 157 | 161 | 166 |
| 0.020 | 0.30 | 101 | 115 | 121 | 132 | 141 | 148 | 153 | 156 | 163 |
| 0.020 | 0.40 | 104 | 114 | 119 | 129 | 139 | 146 | 151 | 154 | 159 |
| 0.020 | 0.50 | 96 | 111 | 117 | 127 | 136 | 143 | 149 | 151 | 158 |
| 0.025 | 0.10 | 107 | 121 | 130 | 139 | 148 | 155 | 162 | 164 | 169 |
| 0.025 | 0.20 | 105 | 116 | 123 | 133 | 143 | 150 | 155 | 158 | 162 |
| 0.025 | 0.30 | 95 | 114 | 120 | 130 | 138 | 146 | 151 | 154 | 159 |
| 0.025 | 0.40 | 94 | 111 | 117 | 127 | 137 | 143 | 149 | 152 | 157 |
| 0.025 | 0.50 | 96 | 108 | 115 | 125 | 134 | 141 | 147 | 150 | 154 |

Table C2: Summary statistics for the simulated earliest time to detection distribution for testing in community surveillance with seedings from 500 daily passengers

| Community survey size | centiles | | | | | | | | |
|---|---|---|---|---|---|---|---|---|---|
| | 1st | 5th | 10th | 25th | 50th | 75th | 90th | 95th | 99th |
| 20000 | 125 | 139 | 147 | 157 | 166 | 174 | 179 | 183 | 189 |
| 30000 | 122 | 137 | 144 | 153 | 163 | 171 | 177 | 180 | 184 |
| 40000 | 124 | 134 | 141 | 152 | 161 | 167 | 173 | 176 | 181 |
| 50000 | 118 | 132 | 139 | 148 | 158 | 166 | 171 | 173 | 180 |
| 60000 | 117 | 131 | 137 | 147 | 157 | 164 | 170 | 173 | 178 |
| 70000 | 114 | 127 | 134 | 145 | 155 | 162 | 168 | 171 | 178 |
| 80000 | 115 | 128 | 135 | 144 | 154 | 161 | 167 | 170 | 176 |
| 90000 | 113 | 126 | 133 | 144 | 153 | 161 | 166 | 169 | 173 |
| 100000 | 112 | 128 | 133 | 144 | 152 | 160 | 165 | 168 | 173 |
| 110000 | 107 | 126 | 131 | 142 | 152 | 158 | 164 | 167 | 172 |
| 120000 | 106 | 123 | 130 | 140 | 150 | 158 | 163 | 167 | 172 |
| 130000 | 107 | 123 | 131 | 141 | 150 | 157 | 164 | 167 | 171 |
| 140000 | 109 | 124 | 130 | 139 | 148 | 156 | 162 | 164 | 169 |
| 150000 | 106 | 122 | 129 | 139 | 148 | 156 | 161 | 164 | 170 |
| 160000 | 107 | 120 | 128 | 139 | 148 | 155 | 160 | 164 | 169 |
| 170000 | 107 | 121 | 128 | 138 | 148 | 155 | 161 | 163 | 169 |
| 180000 | 108 | 119 | 125 | 136 | 146 | 155 | 161 | 164 | 169 |
| 190000 | 106 | 121 | 128 | 137 | 146 | 154 | 159 | 162 | 169 |
| 200000 | 101 | 119 | 126 | 138 | 146 | 153 | 158 | 161 | 166 |

Table B3: Summary statistics for the simulated earliest time to detection distribution for testing at the border with 100 daily passengers, all times refer to days since the index case

| Proportion tested | centiles | | | | | | | | |
|---|---|---|---|---|---|---|---|---|---|
| | 1st | 5th | 10th | 25th | 50th | 75th | 90th | 95th | 99th |
| 0.010 | 118 | 135 | 143 | 159 | 180 | >200 | >200 | >200 | >200 |
| 0.020 | 111 | 131 | 140 | 152 | 170 | 189 | >200 | >200 | >200 |
| 0.030 | 103 | 127 | 134 | 148 | 163 | 179 | 196 | >200 | >200 |
| 0.040 | 108 | 123 | 129 | 142 | 156 | 171 | 186 | 195 | >200 |
| 0.050 | 105 | 118 | 126 | 139 | 153 | 165 | 179 | 188 | >200 |
| 0.060 | 103 | 117 | 125 | 137 | 150 | 163 | 174 | 182 | 199 |
| 0.070 | 96 | 114 | 122 | 135 | 149 | 161 | 172 | 178 | 191 |
| 0.080 | 97 | 114 | 121 | 134 | 146 | 156 | 167 | 172 | 186 |
| 0.090 | 102 | 115 | 122 | 132 | 144 | 156 | 166 | 172 | 184 |
| 0.100 | 101 | 115 | 122 | 132 | 144 | 154 | 163 | 169 | 180 |
| 0.110 | 98 | 112 | 119 | 130 | 141 | 151 | 161 | 167 | 177 |
| 0.120 | 95 | 112 | 117 | 128 | 140 | 150 | 159 | 164 | 174 |
| 0.130 | 91 | 109 | 116 | 127 | 139 | 148 | 157 | 164 | 172 |
| 0.140 | 97 | 111 | 118 | 128 | 138 | 148 | 157 | 161 | 168 |
| 0.150 | 91 | 110 | 117 | 127 | 137 | 146 | 155 | 161 | 169 |
| 0.160 | 91 | 105 | 114 | 126 | 137 | 146 | 154 | 159 | 168 |
| 0.170 | 97 | 110 | 114 | 125 | 135 | 144 | 153 | 158 | 167 |
| 0.180 | 89 | 105 | 114 | 125 | 134 | 143 | 152 | 157 | 164 |
| 0.190 | 91 | 105 | 114 | 124 | 134 | 143 | 151 | 156 | 164 |
| 0.200 | 87 | 104 | 112 | 123 | 133 | 142 | 150 | 154 | 162 |
| 0.210 | 90 | 103 | 112 | 122 | 132 | 142 | 149 | 155 | 161 |
| 0.220 | 89 | 105 | 111 | 122 | 132 | 141 | 148 | 151 | 158 |
| 0.230 | 88 | 104 | 111 | 122 | 133 | 141 | 148 | 153 | 159 |
| 0.240 | 91 | 104 | 110 | 120 | 132 | 141 | 147 | 152 | 159 |
| 0.250 | 88 | 103 | 110 | 120 | 131 | 140 | 148 | 151 | 157 |
| 0.260 | 83 | 103 | 110 | 121 | 131 | 140 | 146 | 150 | 158 |
| 0.270 | 87 | 103 | 110 | 120 | 130 | 139 | 145 | 150 | 156 |
| 0.280 | 80 | 102 | 111 | 120 | 130 | 138 | 144 | 149 | 154 |
| 0.290 | 86 | 101 | 108 | 119 | 129 | 137 | 144 | 148 | 155 |
| 0.300 | 83 | 100 | 107 | 118 | 128 | 137 | 143 | 148 | 156 |
| 0.310 | 90 | 102 | 110 | 119 | 129 | 137 | 143 | 147 | 153 |
| 0.320 | 87 | 103 | 110 | 118 | 128 | 136 | 143 | 147 | 152 |
| 0.330 | 85 | 101 | 108 | 118 | 127 | 136 | 143 | 146 | 154 |
| 0.340 | 88 | 100 | 108 | 117 | 127 | 136 | 142 | 146 | 154 |
| 0.350 | 83 | 101 | 106 | 116 | 126 | 135 | 141 | 146 | 153 |
| 0.360 | 78 | 100 | 107 | 116 | 126 | 135 | 142 | 145 | 152 |
| 0.370 | 85 | 100 | 107 | 117 | 127 | 135 | 141 | 145 | 153 |
| 0.380 | 82 | 97 | 106 | 116 | 126 | 134 | 141 | 145 | 150 |
| 0.390 | 83 | 97 | 104 | 116 | 125 | 134 | 140 | 144 | 152 |
| 0.400 | 78 | 99 | 106 | 116 | 125 | 133 | 139 | 143 | 150 |
| 0.410 | 78 | 98 | 104 | 115 | 125 | 133 | 140 | 143 | 149 |
| 0.420 | 82 | 99 | 106 | 116 | 125 | 133 | 139 | 144 | 151 |
| 0.430 | 82 | 100 | 106 | 116 | 124 | 133 | 138 | 142 | 148 |
| 0.440 | 77 | 100 | 106 | 115 | 124 | 132 | 139 | 142 | 149 |
| 0.450 | 80 | 97 | 104 | 114 | 124 | 133 | 138 | 142 | 148 |
| 0.460 | 81 | 95 | 103 | 115 | 124 | 131 | 138 | 141 | 148 |

| | | | | | | | | | |
|---|---|---|---|---|---|---|---|---|---|
| 0.470 | 87 | 98 | 104 | 114 | 124 | 131 | 138 | 141 | 147 |
| 0.480 | 80 | 97 | 105 | 114 | 123 | 131 | 138 | 141 | 146 |
| 0.490 | 84 | 97 | 104 | 113 | 123 | 131 | 137 | 141 | 149 |
| 0.500 | 84 | 98 | 104 | 113 | 123 | 131 | 136 | 140 | 148 |

Table H3: Summary statistics for the simulated earliest time to detection distribution for testing at hospitals with seedings from 100 daily passengers

| IHR | Proportion tested | centiles | | | | | | | | |
|---|---|---|---|---|---|---|---|---|---|---|
| | | 1st | 5th | 10th | 25th | 50th | 75th | 90th | 95th | 99th |
| 0.005 | 0.10 | 135 | 151 | 159 | 170 | 180 | 189 | 196 | 199 | >200 |
| 0.005 | 0.20 | 125 | 146 | 153 | 164 | 174 | 182 | 188 | 192 | 198 |
| 0.005 | 0.30 | 126 | 141 | 149 | 161 | 171 | 179 | 186 | 189 | 194 |
| 0.005 | 0.40 | 120 | 139 | 146 | 158 | 167 | 175 | 182 | 186 | 192 |
| 0.005 | 0.50 | 118 | 138 | 144 | 156 | 166 | 174 | 180 | 183 | 188 |
| 0.010 | 0.10 | 128 | 146 | 152 | 163 | 173 | 183 | 189 | 192 | 197 |
| 0.010 | 0.20 | 128 | 140 | 147 | 157 | 167 | 176 | 182 | 186 | 190 |
| 0.010 | 0.30 | 120 | 136 | 143 | 153 | 164 | 172 | 178 | 182 | 188 |
| 0.010 | 0.40 | 116 | 134 | 141 | 151 | 161 | 169 | 176 | 180 | 186 |
| 0.010 | 0.50 | 114 | 132 | 140 | 151 | 160 | 168 | 175 | 178 | 184 |
| 0.015 | 0.10 | 126 | 141 | 147 | 159 | 170 | 178 | 185 | 189 | 195 |
| 0.015 | 0.20 | 118 | 135 | 143 | 154 | 164 | 173 | 179 | 182 | 187 |
| 0.015 | 0.30 | 113 | 133 | 140 | 151 | 161 | 169 | 175 | 178 | 185 |
| 0.015 | 0.40 | 112 | 130 | 136 | 147 | 157 | 165 | 173 | 176 | 182 |
| 0.015 | 0.50 | 111 | 130 | 136 | 147 | 156 | 164 | 171 | 174 | 180 |
| 0.020 | 0.10 | 123 | 139 | 147 | 157 | 167 | 176 | 183 | 186 | 193 |
| 0.020 | 0.20 | 118 | 132 | 141 | 151 | 162 | 170 | 176 | 180 | 186 |
| 0.020 | 0.30 | 110 | 129 | 136 | 148 | 157 | 166 | 172 | 176 | 182 |
| 0.020 | 0.40 | 113 | 127 | 134 | 145 | 155 | 164 | 170 | 173 | 179 |
| 0.020 | 0.50 | 106 | 127 | 134 | 144 | 154 | 161 | 168 | 171 | 177 |
| 0.025 | 0.10 | 121 | 136 | 144 | 156 | 165 | 173 | 180 | 183 | 189 |
| 0.025 | 0.20 | 114 | 131 | 138 | 149 | 159 | 167 | 174 | 177 | 182 |
| 0.025 | 0.30 | 114 | 129 | 136 | 146 | 156 | 164 | 171 | 174 | 179 |
| 0.025 | 0.40 | 110 | 126 | 133 | 144 | 153 | 162 | 168 | 171 | 175 |
| 0.025 | 0.50 | 110 | 124 | 131 | 142 | 151 | 159 | 165 | 169 | 173 |

Table C3: Summary statistics for the simulated earliest time to detection distribution for testing in community surveillance with seedings from 100 daily passengers

| Community survey size | centiles | | | | | | | | |
|---|---|---|---|---|---|---|---|---|---|
| | 1st | 5th | 10th | 25th | 50th | 75th | 90th | 95th | 99th |
| 20000 | 139 | 156 | 164 | 175 | 185 | 193 | 200 | >200 | >200 |
| 30000 | 132 | 150 | 158 | 170 | 180 | 189 | 195 | 199 | >200 |
| 40000 | 136 | 151 | 157 | 168 | 178 | 187 | 193 | 197 | >200 |
| 50000 | 128 | 144 | 154 | 165 | 176 | 184 | 191 | 196 | >200 |
| 60000 | 129 | 147 | 153 | 164 | 174 | 183 | 190 | 193 | 198 |
| 70000 | 124 | 144 | 150 | 163 | 173 | 181 | 189 | 192 | 198 |
| 80000 | 125 | 144 | 150 | 162 | 172 | 180 | 186 | 190 | 196 |
| 90000 | 122 | 141 | 149 | 160 | 171 | 179 | 185 | 189 | 194 |
| 100000 | 128 | 142 | 149 | 159 | 169 | 177 | 184 | 187 | 194 |
| 110000 | 123 | 138 | 147 | 158 | 169 | 177 | 183 | 186 | 192 |
| 120000 | 123 | 138 | 147 | 158 | 168 | 176 | 183 | 186 | 192 |
| 130000 | 125 | 140 | 147 | 157 | 167 | 175 | 182 | 185 | 190 |
| 140000 | 124 | 141 | 148 | 158 | 167 | 175 | 182 | 185 | 191 |
| 150000 | 121 | 136 | 144 | 156 | 165 | 174 | 180 | 184 | 191 |
| 160000 | 119 | 138 | 146 | 156 | 165 | 174 | 180 | 184 | 189 |
| 170000 | 116 | 135 | 143 | 153 | 164 | 172 | 179 | 182 | 188 |
| 180000 | 118 | 136 | 143 | 154 | 164 | 173 | 178 | 182 | 188 |
| 190000 | 118 | 135 | 143 | 154 | 164 | 172 | 177 | 181 | 188 |
| 200000 | 117 | 134 | 142 | 153 | 163 | 171 | 178 | 181 | 187 |
